# Reproducibility and Robustness of Large Language Models for Mobility Functional Status Extraction

**DOI:** 10.64898/2026.04.03.26350117

**Authors:** Xingyi Liu, Muskan Garg, Eunji Jeon, Heling Jia, Jennifer St. Sauver, Sandeep R. Pagali, Sunghwan Sohn

## Abstract

Clinical narrative text contains crucial patient information, yet reliable extraction remains challenging due to linguistic variability, documentation habits, and differences across care settings. Large language models (LLMs) have shown strong accuracy on clinical information extraction (IE), but their reproducibility (stability under repeated runs) and robustness (stability under small, natural prompt variations) are less consistently quantified, despite being central to clinical deployment. In this study, we evaluate three open-weight LLMs representing distinct modeling choices: a dense general-purpose model (Llama 3.3), a mixture-of-experts (MoE) general-purpose model (Llama 4), and a domain-tuned medical model (MedGemma). We focus on binary clinical IE aligned with four mobility classes from the International Classification of Functioning, Disability and Health (ICF) framework. Using a controlled experimental design, we quantify (1) intra-prompt reproducibility across repeated sampling and (2) inter-prompt robustness across paraphrased prompts. We jointly report predictive performance (F1-score) and stability (Fleiss’ Kappa). And we test factor effects using three-way ANOVA with post-hoc comparisons. Results show that increasing temperature generally degrades agreement, but the magnitude depends on model and task; furthermore, prompt paraphrasing can substantially reduce stability, with particularly large drops for the MoE model. Finally, we evaluate a practical mitigation, self-consistency via majority voting, which improves κ substantially and often improves or preserves F1-score, at the cost of additional inference. Together, these findings provide a reproducible framework and concrete recommendations for evaluating and improving LLM reliability in clinical IE.

## I. Introduction

Clinical notes contain rich descriptions of patients’ functional status, symptoms, and clinician reasoning, but the information is embedded in heterogeneous and often unstructured narrative text. Large language models (LLMs) have recently shown strong capability for clinical information extraction (IE), sometimes matching or exceeding traditional pipelines on certain tasks and note types. For example, prompt-driven LLM workflows have been evaluated for extracting structured variables from pathology reports at scale, demonstrating promising accuracy while also revealing sensitivity to prompt design and specialized terminology [1].

However, for clinical deployment, accuracy alone is insufficient. Clinical systems require outputs that are stable enough to support downstream analyses, auditing, and end-user trust [2]. Two complementary reliability dimensions are especially relevant in IE settings. The first is reproducibility, meaning the extent to which a model produces consistent extraction decisions when repeatedly queried with the same prompt and the same clinical text. The second is robustness, meaning the extent to which extraction decisions remain stable when the prompt is varied in natural, non-adversarial ways, such as paraphrasing by different clinicians or teams. Empirical evidence from clinical NLP indicates that even plausible changes in instruction wording can produce substantial changes in outputs for clinical classification and extraction tasks, with implications for both reliability and fairness.

Mobility IE is a particularly informative case for studying this sensitivity. Documentation of mobility functional status is sometimes expressed indirectly (e.g., “uses a walker”, “use of floatation for balance”, “no limp”). Mapping these narrative descriptions to standardized mobility categories therefore requires semantic interpretation and reasonable inference, rather than simple keyword matching. As a result, mobility IE provides a useful testbed for evaluating whether context-aware LLM prompting yields not only high average accuracy, but also stable and reproducible extraction decisions under repeated sampling and natural prompt variation.

This paper evaluates reproducibility and robustness in LLM-based clinical IE using a controlled factorial design that varies model family, decoding temperature, and clinical extraction category. We focus on binary IE aligned with mobility classes from the International Classification of Functioning, Disability and Health (ICF) [3], and we compare general-purpose and domain-specific architectures. This comparison is motivated by a deployment-relevant question: whether medical domain adaptation improves not only predictive performance but also reliability, including stability under repeated runs and resilience to instruction rewording. General-purpose models may offer broad linguistic competence, whereas domain-tuned medical models may better reflect clinical documentation conventions and thus yield more stable extraction decisions in practice. Agreement is quantified using Fleiss’ Kappa [4], and predictive performance is reported alongside stability to avoid conflating accuracy with reliability. To understand practical mitigation strategies, we also evaluate self-consistency via majority voting over multiple generations, which has been shown to improve reliability in other LLM settings and provides a straightforward way to reduce stochastic variance without retraining.

## II. Background and Related Work

The task of extracting structured data from unstructured clinical text has long been a central focus of medical informatics. Early approaches relied heavily on rule-based systems (e.g., regex, cTAKES [5]) and traditional Natural Language Processing (NLP) techniques, which, while effective for narrow tasks, struggled with the variability and complexity of clinical language. The advent of transformer-based models like BERT and BioBERT [6] marked a significant leap forward, but these methods typically required extensive, task-specific fine-tuning on large labeled datasets. The current paradigm has shifted toward generative LLMs, which can perform complex information extraction tasks in a zero-shot or few-shot setting through carefully crafted prompts [7]. This approach offers unprecedented flexibility but introduces new challenges. The use of proprietary, API-based architecture raises significant data privacy and security concerns in healthcare, motivating the turn toward open-source models that can be deployed within a secure institutional infrastructure. This study focuses exclusively on open-source models to reflect these practical deployment considerations.

The evaluation of LLM reliability in healthcare is a rapidly growing field. A cross-sectional study used ChatGPT-4 to extract helmet use status of patients injured in micromobility-related accidents from thousands of unstructured clinical notes and found a significant trade-off between prompt simplicity and accuracy [8]. While a highly detailed prompt achieved near-perfect agreement with a rule-based search, less specific prompts yielded only weak to moderate agreement, largely because the LLM failed to correctly identify negations [8]. Critically, their test-retest reliability analysis, using Fleiss’ Kappa, showed that the LLM was not perfectly reproducible and consistently replicated its own hallucinations across sessions.

Sun et al. evaluated multiple LLMs for extracting clinical variables from bladder cancer records and explicitly quantified consistency using Fleiss’ Kappa across repeated runs; they report very high agreement for certain models (including GPT-4 variants) and comparatively lower consistency for others, emphasizing careful model selection and continuous evaluation for clinical use [9].

Prompt-based clinical NLP introduces a deployment-relevant source of distribution shift: instruction text is often authored by different users with different styles. Ceballos-Arroyo et al. conducted a large-scale evaluation of open instruction-tuned models on clinical classification and extraction tasks using prompts written by medical professionals and found substantial performance variability across natural instruction phrasings, including concerning fairness implications under alternative valid prompts [10].

The critical role of prompt design was further explored by Wang et al., who tested various LLMs against evidence-based clinical guidelines using different prompt engineering strategies [11]. They introduced a novel “Reflection of Thoughts” prompt to improve reasoning and found that prompt choice significantly impacted consistency with clinical guidelines. However, they also reported that reliability was unstable, with Fleiss’ Kappa values for repeated queries varying dramatically across models and prompts.

Clinical deployment often requires deterministic or near-deterministic behavior, yet many evaluation settings use stochastic decoding. Temperature is a key parameter controlling output randomness and has been empirically linked to variability in clinically relevant tasks. Jarrett et al. studied temperature-driven variability in emergency diagnostic accuracy with repeated runs and reported systematic degradation in diagnostic accuracy and increased divergence as temperature increases, highlighting the need for transparent reporting and careful parameter selection in clinical AI research [12]. In biomedical text-mining tasks, Windisch et al. examined how temperature affects extraction/classification from clinical trial publications, discussing the lack of consensus on “safe” settings and reporting performance changes at higher temperatures [13]. These findings motivate explicit temperature sweeps when assessing reproducibility in extraction workflows.

Despite prior work on reliability and prompt sensitivity in clinical NLP, a unified assessment that jointly isolates the effects of model family, decoding stochasticity, and instruction rewording on stability for clinical IE remains limited. To fill this gap, our study jointly quantifies reproducibility across repeated runs and robustness across prompt paraphrases under controlled temperature sweeps, and compares general-purpose and domain-specific open-weight architectures on mobility IE.

### III. Experimental Design and Evaluation Framework

#### A. Dataset

This study evaluates LLM reliability on binary clinical IE tasks grounded in the ICF. Four mobility classes are studied:

1. Changing and Maintaining Body Position (ICF codes d410-d429): Encompasses actions such as lying down,sitting, standing, and transferring oneself (e.g., from a bed to a chair).
2. Carrying, Moving and Handling Objects (ICF codes d430-d449): Includes lifting, carrying objects in hands or on the back, and fine hand use like grasping and manipulating objects.
3. Walking and Moving (ICF Changing and Maintaining Body codes d450-d469): Covers walking short or long distances, on different surfaces, and other forms of movement like climbing or running.
4. Moving Around Using Transportation (ICF codes d470-d489): Pertains to using private or public transportation as a passenger or driver.

For each mobility function, 200 annotated clinical note sections were sampled from three Rochester, MN healthcare providers, yielding 800 sections total. The detailed annotation guidelines are provided in the Supplementary Material of [14]. Each function-specific set is well balanced, with half of the sections containing references to the target function and half serving as negatives. For each mobility function, a base prompt was constructed that included the relevant ICF definition and directed the model to output a binary decision (presence or absence). Each model run produced a binary label for each clinical section. For each condition, outputs are represented as a 200-bit vector, where each bit corresponds to a section-level prediction (1 for presence, 0 for absence).

#### B. Models

Three open-weight LLMs were selected to compare general-purpose and domain-specific architectures.

Llama 3.3 70B [15], developed by Meta AI, represents the state of the art in open-weight, dense LLMs as of late 2024. It is built on a standard Transformer architecture employing optimizations like Grouped Query Attention (GQA) to enhance inference efficiency. Pre-trained on a massive corpus of over 15 trillion tokens, it serves as our baseline for a high-capacity “generalist” model. In dense models, every parameter is active for every token generated. This theoretically provides a stable compute path, though the softmax sampling step still introduces randomness.

Llama-4-Scout-17B-16E [16] utilizes a Mixture-of-Experts (MoE) architecture. Unlike dense models, MoEs utilize a “router” or “gating network” to select a sparse subset of experts (e.g., 2 out of 16) to process each token. This allows the model to have a massive total parameter count (109B) while only activating a fraction (17B) for inference, drastically reducing computational cost. However, MoEs introduce a unique source of instability: routing fluctuation. Recent research indicates that the discrete decision-making of the router can be sensitive to minor input perturbations or floating-point variations, leading to different experts being selected for the same input across runs, potentially diverging the generation path.

MedGemma 27B [17], developed by Google, represents the “specialist” approach. Built on the Gemma 3 architecture, it has been specifically adapted for the medical domain through continued pre-training and instruction tuning on a curated mix of public and proprietary medical datasets, including electronic health records (EHRs) and radiology reports.

#### C. Evaluation Metrics

To capture a whole view of model behavior, we evaluate (i) task performance against ground truth and (ii) output stability across repeated generations (reproducibility) and across prompt paraphrases (robustness). In all experiments, each run produces a 200-bit prediction vector over the 200 note sections for a given mobility function.

We emphasize F1-score as the performance metric because it penalizes both false positives and false negatives and provides a single summary of positive-class extraction quality. We report mean performance by averaging metrics across runs within each experimental condition.

To quantify stability of the 200-bit output vectors, we use Fleiss’ Kappa (κ), an inter-rater reliability coefficient for nominal outcomes with more than 3 raters. Here, “raters” correspond to independent model generations.

Let N denote the number of clinical note sections, n the number of raters and *k* the number of categories (*k = 2* for binary classification). Let *n*_*ij*_ be the number of raters who assigned category *j* to clinical note section *i*. The per-section agreement is 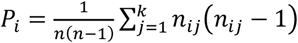. The observed agreement is 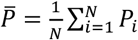. The marginal category proportions are 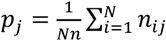 and the chance agreement is 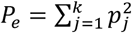 Fleiss’ Kappa κ is then 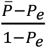 It ranges from 1 to 1, where higher values indicate stronger agreement.

#### D. Experiment Setup

##### a) Experiment 1: Intra-Prompt Reproducibility

To measure reproducibility under identical prompting, the base prompt for each mobility function was executed repeatedly for each model and temperature as shown in Fig. 1. Temperatures were swept across eleven settings from 0.0 to 1.0 in increments of 0.1. The temperature range was selected to target the operational range most commonly used in clinical extraction workflows, where deployment typically prioritizes stable, auditable outputs. This range is sufficient to reveal substantial agreement degradation while keeping decoding within a practically acceptable stochasticity regime. For each model/ function/temperature combination, 100 independent generations were produced. This yields 13,200 total inferences (3 models × 4 functions × 11 temperatures × 100 runs). Reproducibility is quantified by agreement across the 100 output vectors.

**Fig. 1.**
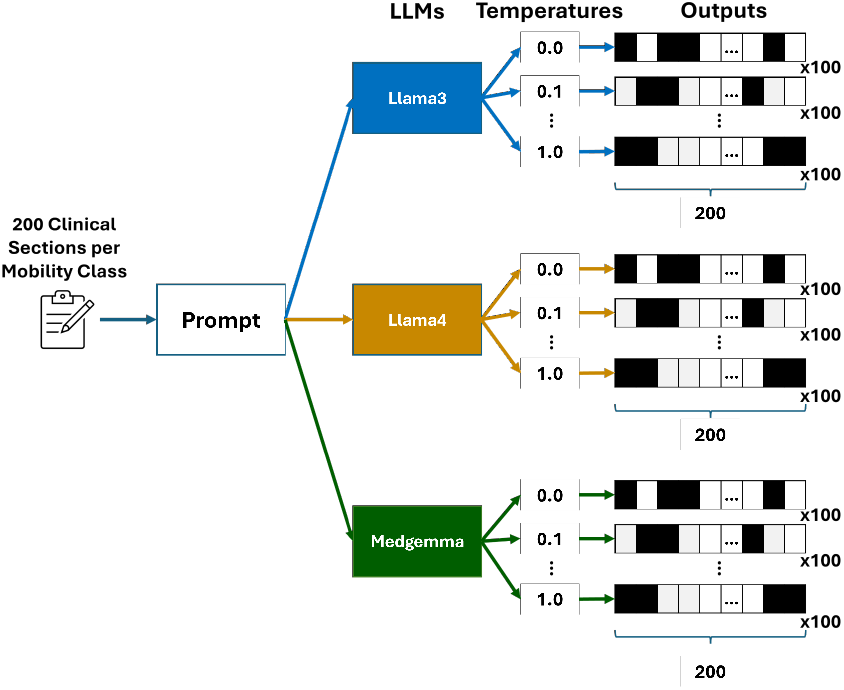
Experiment 1 (intra-prompt reproducibility) design. For each model and mobility class, the base prompt is held fixed and executed across eleven temperature settings from 0.0 to 1.0 in increments of 0.1. At each temperature, the model is run 100 independent times, producing 100 binary prediction vectors over the same 200 clinical note sections. Reproducibility is quantified by computing Fleiss’ Kappa across these repeated runs.

##### b) Experiment 2: Inter-Prompt Robustness

To measure robustness to prompt variation, a family of 10 semantically equivalent paraphrases was manually created for each mobility function as shown in Fig. 2. As summarized in Table I, our prompt paraphrases introduce only surface-form changes while preserving the underlying extraction task. Temperatures were swept across eleven settings from 0.0 to 1.0 in increments of 0.1. For each model/function/paraphrase/temperature combination, the prompt was executed 10 times. We used the 10 repeats to reduce sampling noise within each paraphrase via majority vote, so κ reflects paraphrase sensitivity rather than single-run randomness. This yields 13,200 total inferences (3 models × 4 functions × 10 paraphrases × 11 temperatures × 10 runs). Robustness is quantified by agreement across the paraphrase-conditioned outputs.

**TABLE I.** Representative dimensions of non-adversarial prompt paraphrasing used in Experiment 2, illustrating how semantically equivalent prompts vary.

| Paraphrase variation dimension | Example (Prompt A → Prompt B) |
| --- | --- |
| Instruction framing | “Your task is to determine ...” → “Your objective is to determine ...” |
| Input presentation | “Please analyze the following section ...” → “Inspect the following clinical note section ...” |
| Decision-rule wording | “explicitly or implicitly references ...” → “clearly or subtly indicates ...” |
| Definition header phrasing | “Mobility Category Details” → “Mobility Category Overview” |

**Fig 2.**
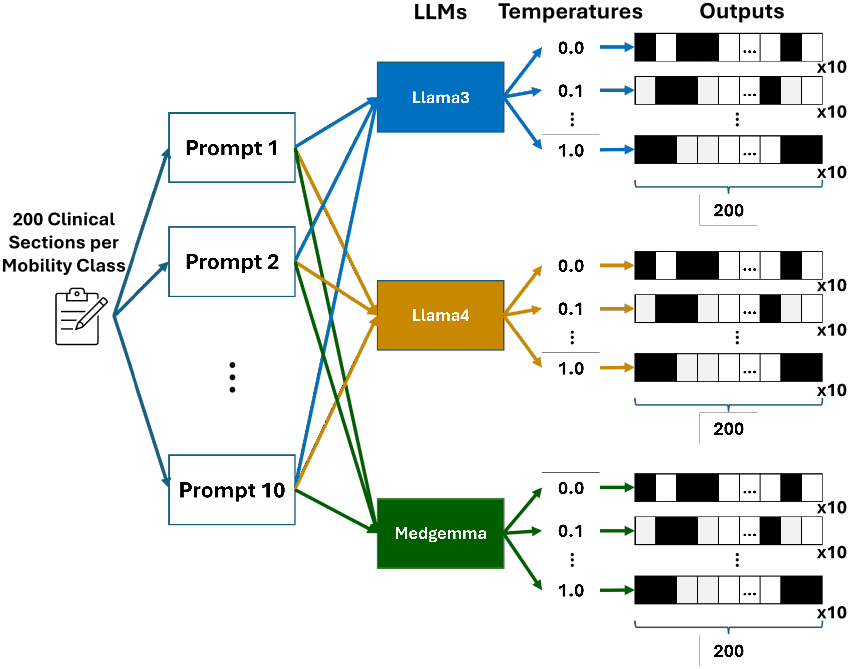
Experiment 2 (inter-prompt robustness) design. For each mobility class, a family of 10 semantically equivalent prompt paraphrases is constructed. For each model and each of eleven temperature settings from 0.0 to 1.0 in increments of 0.1, every paraphrase is executed 10 times, yielding repeated outputs under instruction rewording. Robustness is quantified using Fleiss’ Kappa computed across paraphrase-conditioned outputs.

##### c) Experiment 3: Self-Consistency via Majority Voting

To evaluate a practical mitigation strategy, we implemented self-consistency via majority-vote ensembling. For each condition in Experiment 1, we generated 100 output vectors. These 100 vectors were randomly permuted and partitioned into 10 non-overlapping subsets of size 10. For each subset, we applied a per-section majority vote across the 10 vectors to obtain one aggregated 200-bit prediction vector. This procedure yields 10 ensembled prediction vectors per condition for subsequent evaluation. Agreement (κ) and performance (F1-score) were computed for both single-sample outputs and the ensembled outputs to quantify stability and accuracy changes attributable to self-consistency.

#### E. Statistical Analysis

To test which experimental factors significantly affect stability, separate three-way ANOVAs were conducted for Experiment 1 and Experiment 2. The dependent variable is κ. Factors are Model (3 levels), Temperature (11 levels), and Mobility Class (4 levels), including interaction terms. Where the ANOVA indicated significant main effects, post-hoc Tukey HSD tests were conducted for pairwise model comparisons.

## IV. Results and Analysis

### A. Intra-Prompt Reproducibility Across Temperature

Fig. 3 characterizes intra-prompt behavior under temperature sweeps by jointly plotting mean F1-score (solid) and Fleiss’ κ (dashed), where κ is computed across 100 repeated generations per model/function/temperature setting. Overall, κ is maximal at temperature 0.0 and declines as temperature increases, reflecting the expected increase in sampling stochasticity. However, the rate of degradation is strongly model- and task-dependent: agreement remains comparatively high for “Walking and Moving” and “Moving Around Using Transportation”.

**Fig 3.**
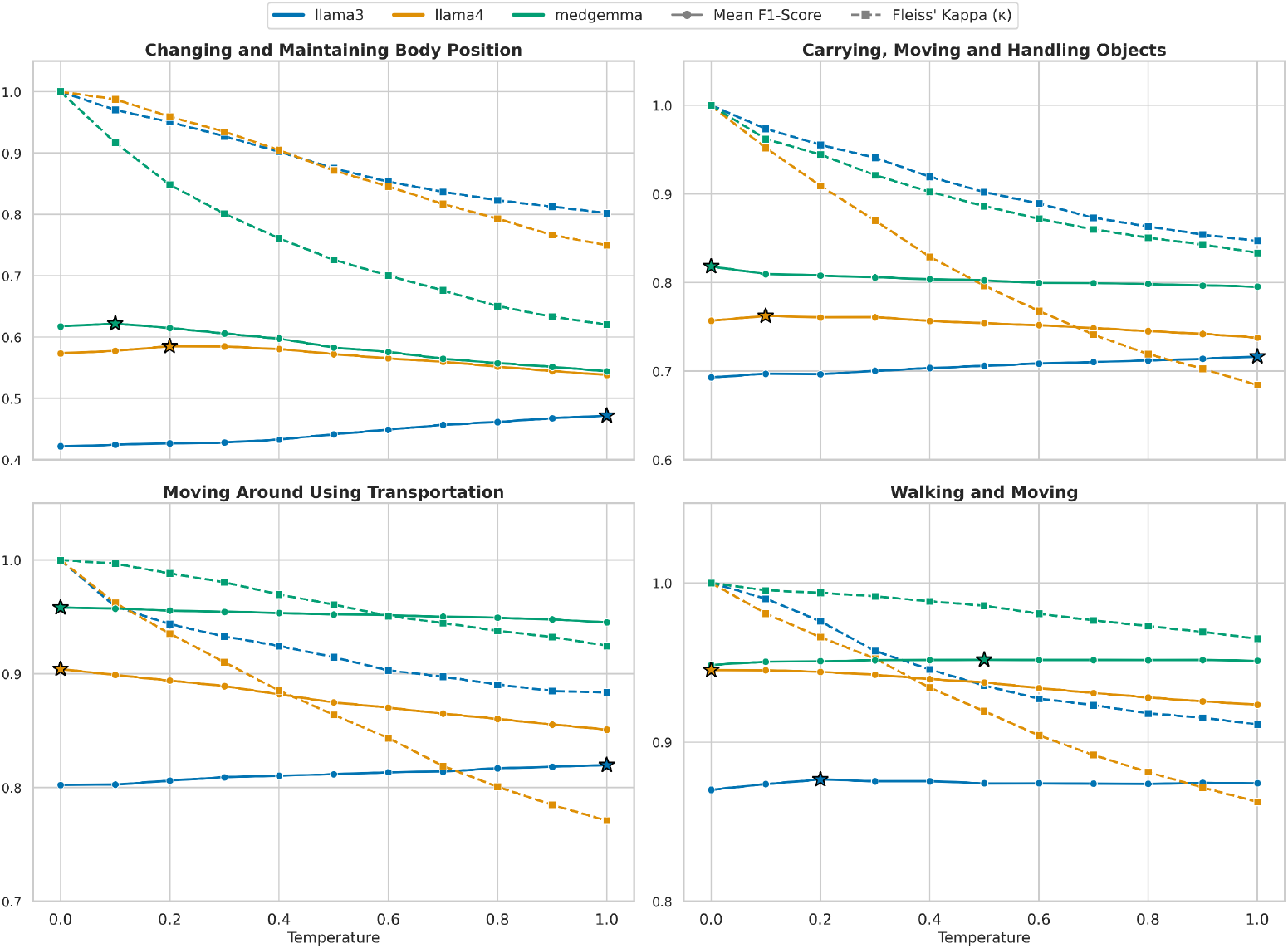
Intra-prompt reproducibility: mean F1-score and Fleiss’ Kappa vs temperature. Intra-prompt reproducibility under temperature sweeps. For each mobility function (four panels), we plot mean F1-score (solid lines) and Fleiss’ Kappa κ (dashed lines) as a function of sampling temperature for three models (Llama 3.3 70B, Llama-4-Scout-17B-16E-Instruct, MedGemma 27B). κ is computed across 100 repeated generations per model/function/temperature combination, quantifying run-to-run stability under an identical prompt. Stars mark the maximum mean F1-score achieved by each model within a mobility function across the temperature sweep.

Importantly, performance trends are substantially flatter than stability trends. Across most model-task pairs, mean F1 changes modestly with temperature, whereas κ can decrease substantially, indicating that average task performance can mask clinically relevant run-to-run variability. In Fig. 3, stars mark the maximum mean F1-score achieved by each model within a mobility function across the temperature sweep, highlighting that the temperature that optimizes performance for a given model is not necessarily the temperature that maximizes reproducibility.

Model-specific behavior is consistent across the four mobility functions. Llama 3.3 shows the most gradual κ decay with increasing temperature, and in several tasks its mean F1-score exhibits a slight upward drift at higher temperatures. It may occur when limited stochasticity helps the model avoid consistent but suboptimal deterministic decisions. Llama 4 demonstrates the steepest κ degradation across temperatures, particularly in “Carrying, Moving and Handling Objects” and “Moving Around Using Transportation”, indicating that stochastic decoding can amplify run-to-run variability for this architecture even when mean F1-score remains competitive. MedGemma shows notable task dependence: it maintains strong performance across the mobility tasks but exhibits a sharper κ drop for “Changing and Maintaining Body Position” as temperature increases, while remaining comparatively stable for “Walking and Moving” and “Moving Around Using Transportation”.

From a deployment perspective, these patterns support conservative decoding choices when repeatability is required. For Llama 3.3, a practical default is to keep temperature at 0.0 for deterministic extraction; this yields maximal reproducibility and avoids run-to-run drift, while the modest F1-score gains observed at higher temperatures in some mobility classes do not typically justify the stability loss in clinical pipelines. In contrast, Llama 4 and MedGemma both achieve their highest mean F1-score at lower temperatures, meaning that increasing temperature provides little or no performance benefit while consistently reducing agreement. Accordingly, for both models, temperature 0.0 is recommended as the operational default for clinical information extraction. Given that MedGemma combines consistently strong performance with high reproducibility at low temperature across all four mobility classes, MedGemma at temperature 0.0 is a reasonable single-configuration choice for deployment when a uniform setting across mobility functions is preferred.

### B. Inter-Prompt Robustness to Paraphrasing

Fig. 4 reports inter-prompt robustness by measuring agreement (Fleiss’ Kappa; dashed lines) across a family of semantically equivalent prompt paraphrases, while simultaneously tracking mean F1-score (solid lines) across the same temperature sweep. Overall, κ values are consistently lower than in the intra-prompt setting, confirming that rewordings of an instruction can induce substantial variability in clinical extraction outputs. As temperature increases, κ generally declines further, but the strength of this effect depends strongly on both the model and the mobility class, indicating that robustness is not a uniform property of a model.

**Fig 4.**
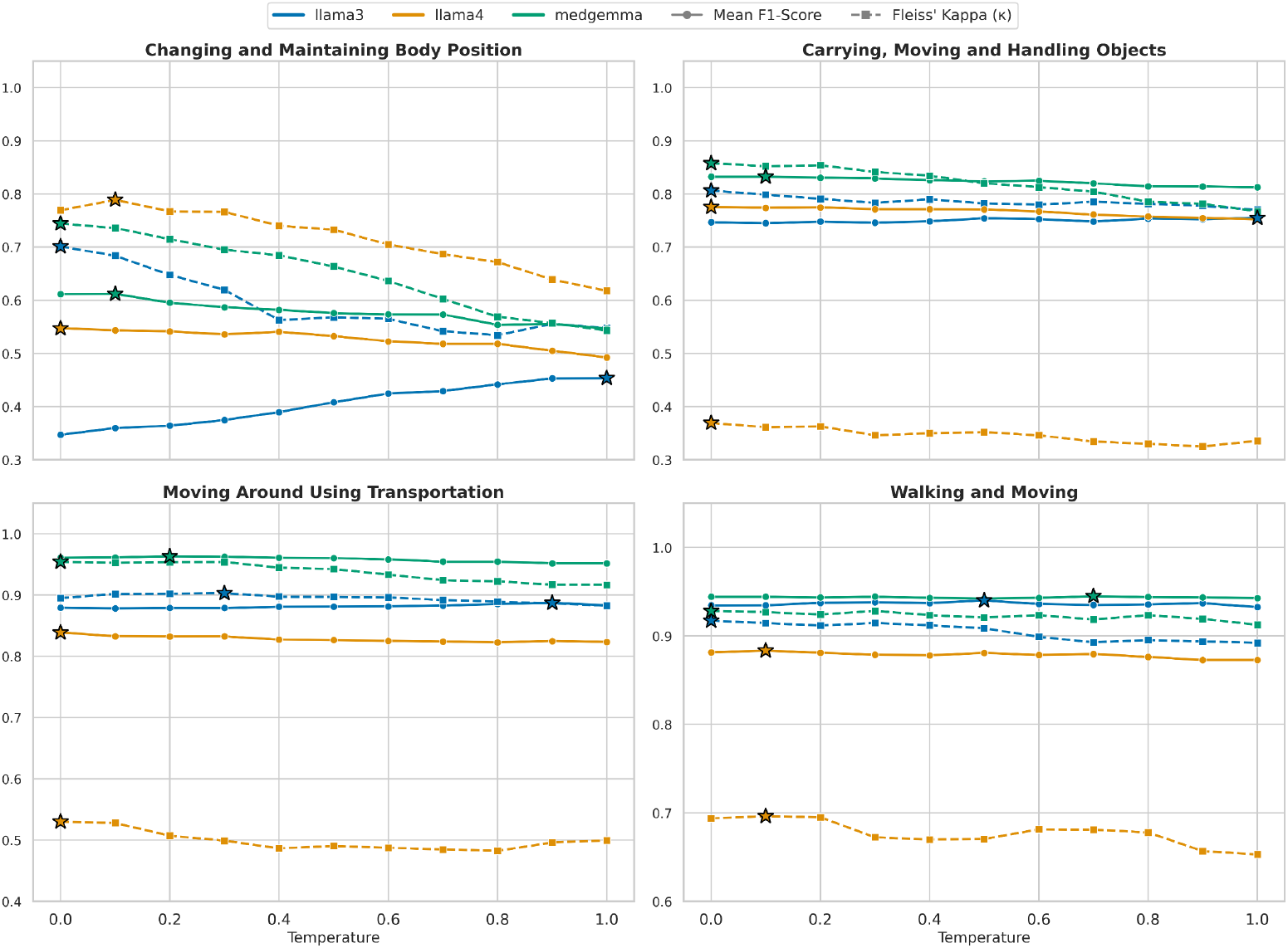
Inter-prompt robustness to semantic paraphrasing under temperature sweeps. For each mobility function (four panels), we plot mean F1-score (solid lines) and Fleiss’ Kappa κ (dashed lines) across sampling temperatures for three models. Here, κ is computed across the 10 prompt paraphrases (treated as raters) for each model/function/temperature combination, measuring stability to instruction rewording. Stars mark the maximum mean F1-score or κ achieved by each model within a mobility function across the temperature sweep.

A key finding is the clear model separation in robustness. Llama 3.3 and MedGemma remain comparatively robust to paraphrasing across most tasks, with κ typically staying high and decreasing only modestly with temperature. In contrast, Llama 4 shows markedly lower robustness in three of the four mobility classes. The degradation is especially striking for “Carrying, Moving and Handling Objects”, where κ remains extremely low across all temperatures, and for “Moving Around Using Transportation”, where κ is also substantially below the other models. Notably, Llama 4 behaves differently on “Changing and Maintaining Body Position”, where its κ is comparatively higher than in its other tasks, underscoring that prompt sensitivity can be highly task-specific even within the same model.

Performance trends again diverge from stability trends. Mean F1-score varies little with temperature for most model-task pairs. For Llama 4 and MedGemma, the highest F1-scores occur at lower temperatures, meaning that increasing temperature provides little performance benefit while consistently worsening robustness. Llama 3.3 is the main exception, where mean F1-score improves at higher temperatures in three of the four mobility classes.

These observations motivate deployment-oriented guidance for paraphrase-prone settings (e.g., multiple clinicians or teams authoring prompts). For Llama 3.3, temperature 0.0 is the safest default when consistent behavior across prompt variants is required; if higher temperature is used to recover performance on specific classes, it should be accompanied by explicit robustness validation across a representative paraphrase set. For Llama 4 and MedGemma, the data support keeping temperature at 0.0 as the operational default because both achieve their best F1 at low temperatures and lose robustness as temperature increases. In particular, given MedGemma’s consistently strong F1-score and comparatively high κ across mobility classes, MedGemma at temperature 0.0 is a practical single-configuration choice when a deployment must tolerate natural prompt rewording while maintaining stable extraction behavior.

### c. Factor Effects on Stability

To quantify which experimental factors drive output stability, we conducted separate three-way ANOVAs on Fleiss’ Kappa for Experiment 1 (intra-prompt reproducibility) and Experiment 2 (inter-prompt robustness to paraphrasing). The ANOVA summaries are reported in Table II.

For reproducibility, we observe significant main effects of Model, Temperature, and Mobility Class, as well as significant interaction effects (Model×Temperature, Model×Mobility Class, and Temperature×Mobility Class). These results indicate that stability is not determined by temperature alone: the extent of κ degradation under stochastic decoding depends on the model architecture and varies across mobility categories.

**TABLE II.** Three-way ANOVA summary for Experiment 1 (intra-prompt reproducibility) and Experiment 2 (inter-prompt robustness to paraphrasing. Factors are Model (M), Temperature (T), and Mobility Class (C); interaction terms are denoted by “×”. *df*_1_ and *df*_2_are effect and residual degrees of freedom, respectively; F is the ANOVA F-statistic, and *p* denotes the corresponding *p*-value. Larger F indicates a larger ratio of explained variance to residual variance, while smaller p indicates stronger statistical evidence against the null hypothesis of no effect.

| Effect | $df_1$ | $df_2$ | Exp. 1: Reproducibility | | Exp. 2: Robustness | |
| --- | --- | --- | --- | --- | --- | --- |
| | | | F | $p$ | F | $p$ |
| M | 2 | 60 | 39.02 | $1.39 \times 10^{-11}$ | 4622.40 | $1.92 \times 10^{-66}$ |
| T | 10 | 60 | 73.40 | $7.68 \times 10^{-30}$ | 34.44 | $3.44 \times 10^{-21}$ |
| C | 3 | 60 | 135.44 | $1.13 \times 10^{-26}$ | 1348.42 | $5.47 \times 10^{-55}$ |
| M × T | 20 | 60 | 2.12 | $1.32 \times 10^{-2}$ | 1.13 | $3.49 \times 10^{-1}$ |
| M × C | 6 | 60 | 55.56 | $1.14 \times 10^{-22}$ | 1121.50 | $1.20 \times 10^{-59}$ |
| T × C | 30 | 60 | 3.99 | $2.55 \times 10^{-6}$ | 8.79 | $7.98 \times 10^{-13}$ |

For robustness under paraphrased prompts, Table II again shows significant main effects of Model, Temperature, and Mobility Class, confirming that prompt rewording introduces substantial variability that differs systematically across models and tasks. Unlike the reproducibility setting, the Model×Temperature interaction is not significant, suggesting that model differences in robustness are broadly persistent across temperatures rather than emerging only at high stochasticity. At the same time, the significant Model×Mobility Class and Temperature×Mobility Class interactions indicate strong task dependence: some mobility classes are more sensitive to paraphrasing and/or temperature than others.

To determine which models differ significantly in overall stability, we performed Tukey HSD post-hoc tests on the main effect of Model for each experiment (Table III). For intra-prompt reproducibility, Llama 3.3 exhibits significantly higher agreement than Llama 4 (*p*=0.0463), while MedGemma does not differ significantly from either model at α= 0.05. For inter-prompt robustness, the post-hoc results show a clearer separation: Llama 4 differs significantly from both Llama 3.3 and MedGemma (both *p*<0.001), whereas Llama 3.3 and MedGemma are not significantly different. This statistical pattern aligns with the trends in Fig. 4, where the MoE model exhibits substantially lower κ under paraphrased prompting, and supports the conclusion that prompt robustness is a distinguishing property of model choice in this clinical IE setting.

**TABLE III.** HSD post-hoc tests for the main effect of Model on Fleiss’ kappa. Here, M1 denotes Model 1 and M2 denotes Model 2 ; L3, L4, and M refer to Llama 3.3, Llama 4, and MedGemma, respectively. The meandiff column reports the difference in mean agreement and *p*_ADJ_ denotes the family-wise error rate (FWER) adjusted p-value from Tukey’s HSD. Negative meandiff indicates higher mean k for M1, positive meandiff indicates higher mean k for M2, and a larger absolute meandiff indicates a larger difference in agreement. The 95% CI provides the uncertainty range for meandiff; intervals that do not include 0 correspond to statistically significant differences at the reported *p*_ADJ_ level.

| Experiment | M1 | M2 | meandiff | $p_{\text{adj}}$ | 95% CI (lower) | 95% CI (upper) |
| --- | --- | --- | --- | --- | --- | --- |
| Exp. 1: Reproducibility | L3 | L4 | -0.0446 | 0.0463 | -0.0887 | -0.0006 |
|  | L3 | M | -0.0157 | 0.6750 | -0.0598 | 0.0283 |
|  | L4 | M | 0.0289 | 0.2687 | -0.0151 | 0.0729 |
| Exp. 2: Robustness | L3 | L4 | -0.2348 | <0.001 | -0.3034 | -0.1661 |
|  | L3 | M | 0.0376 | 0.3984 | -0.0310 | 0.1062 |
|  | L4 | M | 0.2724 | <0.001 | 0.2037 | 0.3410 |

#### D. Effect of Self-Consistency (Majority Voting)

To evaluate whether simple self-consistency can mitigate decoding variability without retraining, we apply majority voting over repeated generations at each temperature and compare the resulting ensembled predictions to the original single-run outputs. Fig. 5 reports mean F1-score under the two settings, and Fig. 6 reports the corresponding Fleiss’ Kappa.

**Fig. 5.**
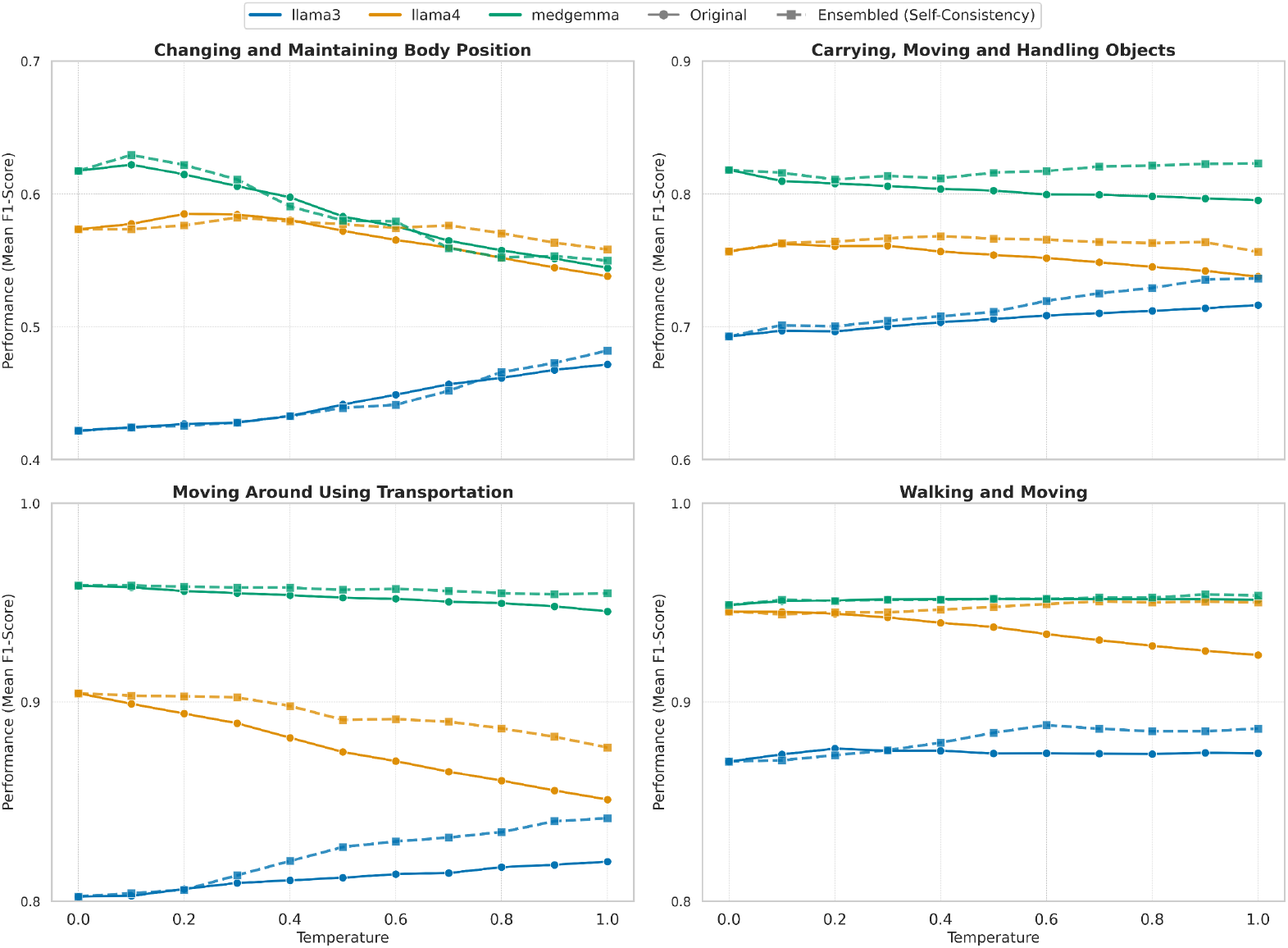
Effect of self-consistency (majority voting) on intra-prompt performance. Mean F1-score versus temperature for each mobility class (four panels), comparing single-run predictions (“Original”) to majority-vote ensembling (“Ensembled”). For each model/function/temperature combination, the ensembled outputs are formed by partitioning the 100 runs from Experiment 1 into 10 groups of 10 generations and taking a per-section majority vote within each group to produce 10 aggregated prediction vectors; F1-score is then averaged over these aggregated vectors.

**Fig. 6.**
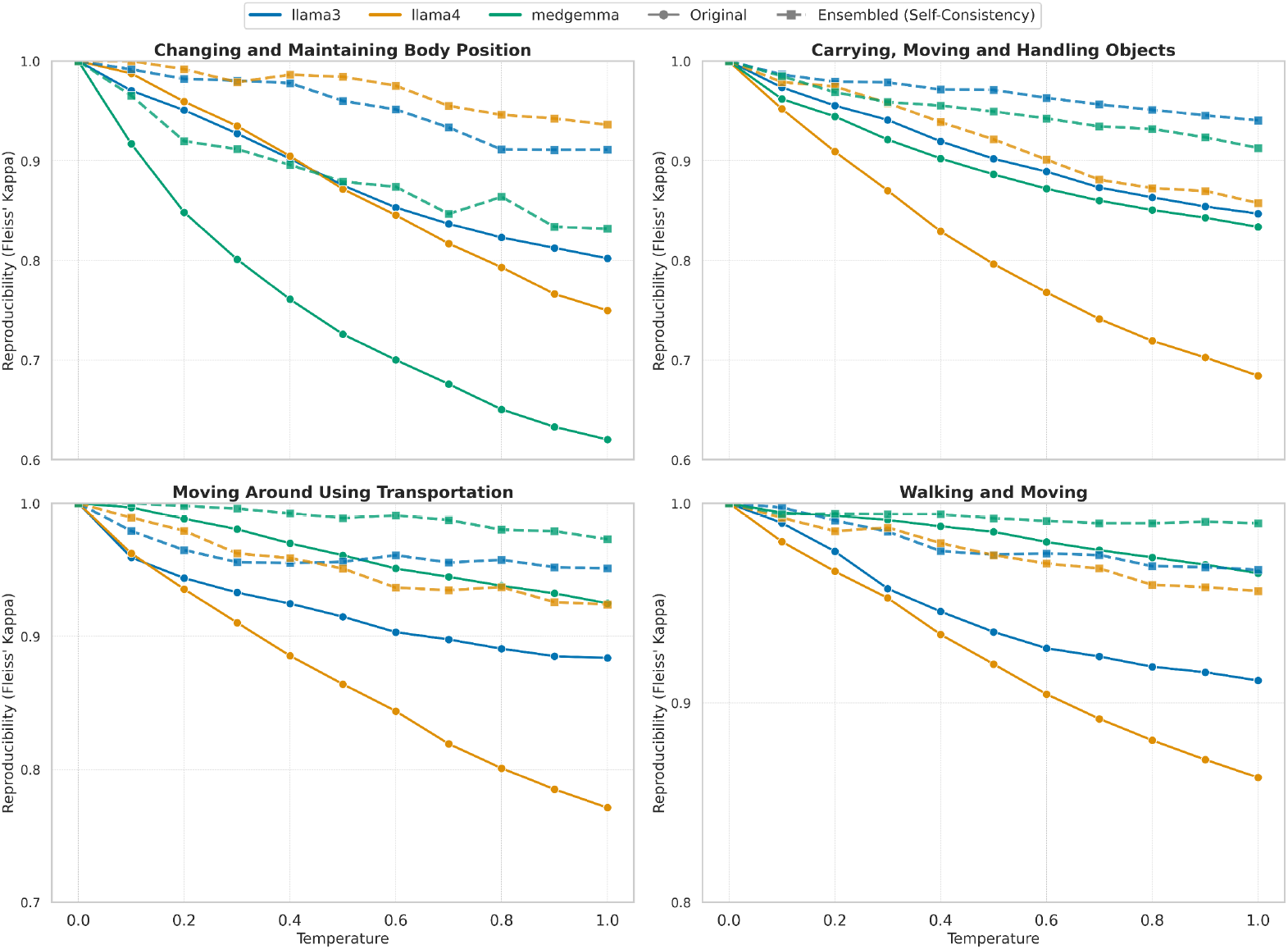
Effect of self-consistency (majority voting) on intra-prompt reproducibility. Fleiss’ Kappa (κ) versus temperature for each mobility class (four panels), comparing reproducibility from single-run outputs (“Original”) to reproducibility after majority-vote ensembling (“Ensembled”). For each model/function/temperature combination, the ensembled outputs are formed by partitioning the 100 runs from Experiment 1 into 10 groups of 10 generations and taking a per-section majority vote within each group to produce 10 aggregated prediction vectors. κ is then computed across the aggregated vectors.

Across all mobility classes, majority voting produces modest but consistent changes in mean F1-score. For Llama 3.3, the original (single-run) mean F1-score increases as temperature rises across multiple mobility classes. Majority voting preserves and typically amplifies this upward trend, yielding consistently higher F1 than the original curve at the same temperature, particularly at moderate-to-high temperatures where stochastic decoding appears to provide marginal accuracy gains. For Llama 4, ensembling primarily acts as a stabilizer against temperature-induced performance drift: the ensembled curves remain comparatively flat with temperature and partially recover the downward trend observed in the original outputs, especially for “Moving Around Using Transportation” and “Walking and Moving”. MedGemma shows relatively small F1 changes because its baseline performance is already strong and stable across temperatures in most mobility classes; ensembling provides incremental gains or preserves performance rather than notably shifting it.

The impact on reproducibility is substantially larger than the impact on F1-score. In every mobility class and for all three models, self-consistency shifts κ upward and flattens the κ-temperature relationship, indicating reduced sensitivity to stochastic decoding. The effect is most significant for Llama 4, which exhibits steep κ degradation at higher temperatures in the original setting; majority voting restores high agreement and maintains κ at a substantially higher level across the full temperature range. Llama 3.3 also benefits strongly while its original κ declines with temperature, the ensembled κ remains consistently higher and decays more slowly. For MedGemma, the benefit is task-dependent but still meaningful, most notably for “Changing and Maintaining Body Position”.

Practically, these results suggest that self-consistency is an effective reliability intervention when repeated calls are feasible. It delivers large reproducibility gains (and occasional performance improvements) while requiring only inference-time changes. The tradeoff is cost since majority voting increases compute and latency roughly in proportion to the number of samples. In deployment settings where deterministic behavior is preferred, temperature 0.0 remains the simplest and most cost-effective choice. When nonzero temperatures are unavoidable (e.g., integration constraints, multi-agent systems, or exploration-oriented prompting), majority voting provides a robust safeguard.

## V. Discussion

This study quantifies the reliability of LLM-based clinical IE along two deployment-relevant dimensions: intra-prompt reproducibility and inter-prompt robustness. Across both experiments, a central observation is that accuracy alone is not a sufficient indicator of reliability. In multiple model-task conditions, mean F1-score varied only modestly across temperatures while agreement declined substantially, implying that aggregate performance can obscure clinically meaningful run-to-run variability. This distinction matters in downstream clinical NLP pipelines where small instabilities can translate into inconsistent inclusion criteria, irreproducible analytics, and reduced auditability.

A second key result is that benign prompt paraphrasing can meaningfully reduce stability. Even when prompts are semantically equivalent, small wording differences produce large agreement drops, with effect sizes that depend strongly on both the model and the clinical function. Notably, the MoE model exhibits particularly low prompt robustness across multiple tasks, and statistical testing confirms model as the dominant determinant of robustness, with these differences persisting across temperatures. Practically, this suggests that prompt standardization alone may be insufficient: model selection should explicitly account for prompt sensitivity in settings where prompts are authored by multiple stakeholders or evolve over time.

Model comparisons highlight architecture-dependent reliability behavior. The dense general-purpose model exhibited comparatively gradual stability decay with increasing temperature, whereas the MoE model showed markedly lower robustness under paraphrasing in several mobility classes, consistent with the broader concern that routing behavior may introduce additional variance beyond sampling stochasticity. The domain-tuned medical model combined strong predictive performance with relatively high stability at lower temperatures across mobility tasks, suggesting that medical adaptation may help reduce sensitivity in this clinical IE setting. Together, these patterns reinforce that reliability is not a single global attribute of “using an LLM”, but an empirical property of a specific model/task/decoding configuration that should be measured rather than assumed.

Finally, the self-consistency results indicate that majority voting is a practical, model-agnostic mitigation. Ensembling consistently improved agreement and often preserved or modestly improved performance, with the largest stability gains occurring in conditions that were most variable under single-run sampling. This tradeoff is operationally important: majority voting can be viewed as an inference-time reliability control that reduces variance without retraining, but it increases latency and compute roughly proportionally to the number of samples. As a result, deployment decisions should explicitly balance reliability requirements against cost constraints. In settings where deterministic outputs are required and throughput is critical, conservative decoding (e.g., temperature near 0) remains the simplest approach; where non-determinism is unavoidable or where higher-temperature decoding is used to recover performance, self-consistency offers a straightforward safeguard.

## VI. Limitations and Future Work

This study has several limitations that motivate follow-on work. First, the evaluation focuses on four binary mobility categories aligned with ICF definitions. While these tasks are clinically meaningful and representative of functional status extraction, findings may not directly generalize to other IE settings such as span-level extraction, relation extraction, temporal reasoning, or multi-label phenotyping. Extending this framework to additional clinical tasks and output structures would clarify how reproducibility and robustness behave under higher linguistic and inferential complexity.

Second, the dataset consists of annotated note sections drawn from a limited set of providers within a single health system. Institutional documentation style, note templates, and patient populations may influence both performance and stability. Multi-site validation and cross-institutional replication are important next steps to establish external validity and to understand whether prompt sensitivity differs across documentation regimes.

Third, robustness was assessed using ten curated paraphrases per task. Real-world prompt variation may be broader and may include formatting changes, different output schemas, multi-turn context, tool-augmented prompting, and templated instructions embedded within clinical workflows. Future work should evaluate robustness under richer perturbation families and quantify how “prompt drift” over time affects longitudinal reproducibility in deployed systems.

Fourth, this work centers on temperature as the primary decoding control. Other decoding parameters (e.g., top-p, top-k, repetition penalties) and system-level factors (e.g., context window length, batching, hardware variability) may also affect stability. A more comprehensive decoding analysis, including deterministic constrained decoding and structured output enforcement, could identify configurations that better preserve both reliability and accuracy for clinical IE. We did not evaluate temperatures greater than 1; future work can extend the sweep to characterize higher-stochasticity regimes.

Future studies should examine which error modes (false positives or false negatives) are most sensitive to paraphrasing and sampling variability. Beyond evaluation, a key future direction is reliability-oriented optimization, including stability-aware fine-tuning, calibration, paraphrase-invariant prompting strategies, and methods tailored to MoE routing stability, with the goal of improving robustness without requiring large ensembles at inference time.

## VII. Conclusion

This work presents a controlled, statistically grounded evaluation of reproducibility and robustness for LLM-based clinical information extraction, using open-weight models that span dense general-purpose, MoE general-purpose, and domain-tuned medical architectures. Avoiding reliance on accuracy alone, we jointly report mean F1-score and Fleiss’ Kappa to characterize both predictive quality and stability under systematic temperature sweeps and under natural prompt paraphrasing. Results demonstrate that stability can degrade substantially, paraphrasing can introduce meaningful output variability, and these effects depend strongly on the interaction of model architecture and clinical category. We further show that self-consistency via majority voting provides a practical inference-time mechanism to improve agreement across models and tasks. Collectively, these findings support routine reporting of stability metrics alongside performance and motivate reliability-focused evaluation for safe and auditable clinical deployment of LLM-based extraction systems.

## Data Availability

All data used for this work was from electronic health records which include identifiable data and thus cannot be made publicly available due to privacy and legal reasons.

## Acknowledgment

This study was supported by Eric and Wendy Schmidt Fund for AI Research and Innovation, NIH (National Institutes of Health) R01 AG068007 and RF1 AG090341. The content is solely the responsibility of the authors and does not necessarily represent the official views of the National Institutes of Health.

